# Tissue-specific tolerance in fatal Covid-19

**DOI:** 10.1101/2020.07.02.20145003

**Authors:** David A Dorward, Clark D Russell, In Hwa Um, Mustafa Elshani, Stuart D Armstrong, Rebekah Penrice-Randal, Tracey Millar, Chris EB Lerpiniere, Giulia Tagliavini, Catherine S Hartley, Nadine P Randle, Naomi N Gachanja, Philippe MD Potey, Alison M Anderson, Victoria L Campbell, Alasdair J Duguid, Wael Al Qsous, Ralph BouHaidar, J Kenneth Baillie, Kevin Dhaliwal, William A Wallace, Christopher OC Bellamy, Sandrine Prost, Colin Smith, Julian A Hiscox, David J Harrison, Christopher D Lucas, on behalf of the ICECAP consortium

## Abstract

**Background:** Tissue inflammation is associated with organ dysfunction and death in Covid-19. The efficacy of dexamethasone in preventing mortality in critical Covid-19 suggests that inflammation has a causal role in death. Whether this deleterious inflammation is a direct response to the presence of SARS-CoV-2, or an independent immuno-pathologic process, is unknown.

**Methods:** Tissue was acquired from detailed post-mortem examinations conducted on 11 well characterised hospitalised patients with fatal Covid-19. SARS-CoV-2 organotropism was mapped at an organ level by multiplex PCR and sequencing, with cellular resolution achieved by in situ viral spike (S) protein detection. Histological evidence of inflammation and organ injury was systematically examined, and the pulmonary immune response characterized with multiplex immunofluorescence.

**Findings:** SARS-CoV-2 was detected across a wide variety of organs, most frequently in the respiratory tract but also in numerous extra-pulmonary sites. Minimal histological evidence of inflammation was identified in non-pulmonary organs despite frequent detection of viral RNA and protein. At a cellular level, viral protein was identified without adjacent inflammation in the intestine, liver and kidney. Severe inflammatory change was restricted to the lung and reticulo-endothelial system. Diffuse alveolar damage, pulmonary thrombi and a monocyte/myeloid-predominant vasculitis were the predominant pulmonary findings, though there was not a consistent association between viral presence and either the presence or nature of the inflammatory response within the lung. Immunophenotyping revealed an influx of macrophages, monocytes and T cells into pulmonary parenchyma. Bone marrow examination revealed plasmacytosis, erythroid dysplasia and iron-laden macrophages. Plasma cell excess was also present in lymph node, spleen and lung. These stereotyped reticulo-endothelial responses occurred largely independently of the presence of virus in lymphoid tissues.

**Conclusions:** Tissue inflammation and organ dysfunction in fatal Covid-19 do not map to the tissue and cellular distribution of SARS-CoV-2, demonstrating tissue-specific tolerance. We conclude that death in Covid-19 is primarily a consequence of immune-mediated, rather than pathogen-mediated, organ inflammation and injury.

**Funding:** The Chief Scientist Office, LifeArc, Medical Research Scotland, UKRI (MRC).

## INTRODUCTION

Inflammation, organ injury and death due to viral infection can occur as a result of direct viral cytotoxicity, collateral damage from an appropriate pathogen-driven immune response, or an aberrant response precipitated by the pathogen, causing immunopathology. Resilience to infectious disease is frequently thought of as best achieved through resistance (controlling pathogen load to prevent organ injury) but the emerging concept of pathogen tolerance (preventing organ injury and inflammation despite the presence of virus) is equally valid.^1^ In this context tolerance could involve restricting the production of injurious inflammatory effectors or moderating pro-inflammatory signalling, downstream of pathogen sensing^2,6^.

Hyper-inflammation is a recognised component of Covid-19, associated with organ dysfunction and death.^3,4^ Fatal Covid-19 most often occurs due to critical impairment of oxygenation and treatment with the corticosteroid dexamethasone reduces mortality in these circumstances.^5,7–10^ This suggests that pulmonary inflammation is causal in death and promoting tolerance may be beneficial, but it remains unknown whether this inflammation is a direct response to the presence of SARS-CoV-2 or an independent immunopathologic process. While Covid-19 is principally thought of as a pulmonary disease, emerging data shows that SARS-CoV-2 also has extra-pulmonary tissue tropism.^11^ However, systematic correlation with histological and clinical evidence of organ injury/inflammation at a tissue and cellular level remains poorly defined.

In order to better understand the pathogen-host interaction in Covid-19, we performed detailed autopsies to present a multi-parameter tissue survey of fatal Covid-19. We sought to characterise viral organotropism and organ-specific inflammatory responses, and to determine the relationships between the presence of virus, local and systemic inflammation, and evidence of organ injury throughout the body.

## RESULTS

### Mapping SARS-CoV-2 distribution to tissue inflammation

To create a detailed tissue atlas of fatal Covid-19 we sampled 37 distinct anatomical tissue sites to identify viral RNA distribution and host inflammatory responses. We detected SARS-CoV-2 RNA across all sampled organs and tissue sites, most frequently in the respiratory tract but also from the gastrointestinal tract, heart and muscle, and less often from the liver, kidney and other organs (Fig. 1a,b). Despite all sampled organs having the potential to contain SARS-CoV-2 RNA, we observed substantial inter-patient variation in the tissue sites involved (Fig. 1b). The time from illness onset to death did not correlate with the number of PCR-positive organs (Fig. 1b). Results from multiplex PCR were confirmed to map to the SARS-CoV-2 genome by sequencing, significantly increasing confidence in these data compared with a PCR only approach.

**Figure 1.**
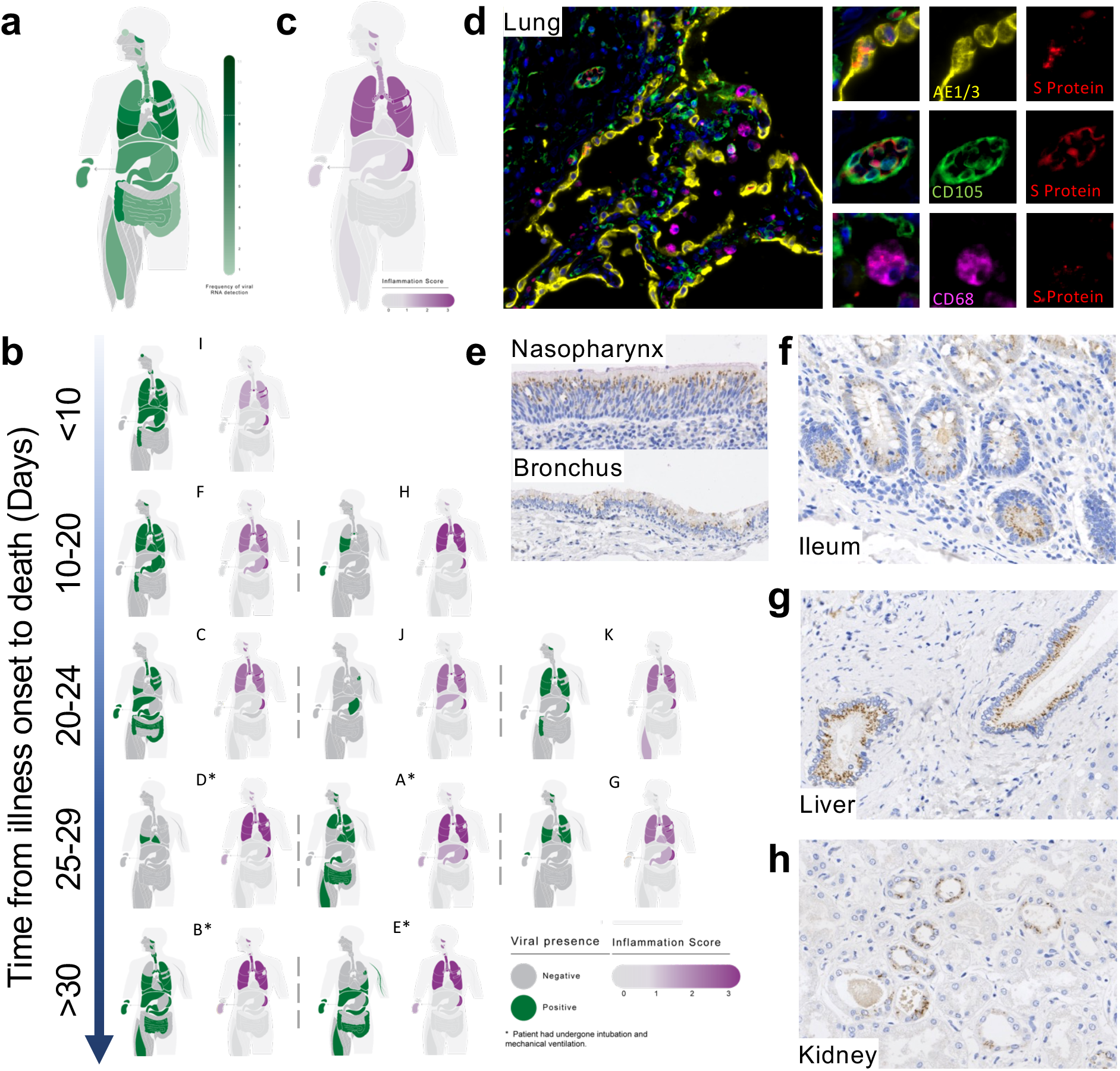
Mapping SARS-CoV-2 organotropism and cellular distribution in fatal Covid-19 in relation to local inflammation. Distribution of SARS-CoV-2 RNA for all patients was determined by multiplex PCR (A; colour intensity denotes frequency of detectable RNA, dotted line on legend denotes maximal frequency within the patient cohort) (n=11). Distribution of individual patient viral RNA presence within organs plotted against time interval between illness onset and death compared with semi-quantitative score of organ specific inflammation for each patient (B). Severity of acute organ injury was assessed semi-quantitatively (0-3; no acute change (0) to severe organ injury/histological abnormality (3)) with aggregate scores visualised (C; n=10-11 per organ/tissue site). Cellular distribution of SARS-CoV-2 S protein was evaluated by immunohistochemistry and multiplex immunofluorescence on FFPE tissue demonstrating its presence within alveolar epithelium and rarely in macrophages and endothelium within the lung parenchyma (D) nasal mucosal and bronchial epithelium (E), as well as small bowel enterocytes (F), distal biliary epithelium within the liver (G) and distal renal tubular epithelium (H). Representative images from n=4 PCR-positive patients.

As analysis of SARS-CoV-2 RNA confirmed presence in numerous organs, detailed histological analysis of multiple tissues was undertaken on every patient to determine the associated pathological consequences and inflammatory responses. In contrast to the distribution of viral RNA, this analysis indicated that the lung and reticulo-endothelial system were the exclusive sites of an extensive inflammatory response (Fig. 1b,c). Extra-pulmonary sites of viral presence did not have substantial local inflammation.

To better resolve the organ-specific pathogen-host interaction at a spatial and cellular level, the presence of S protein was evaluated on select SARS-CoV-2 PCR-positive tissues. Consistent with the latest reports on tissue expression of SARS-CoV-2 entry factors^12^, S protein was found predominantly within epithelia of the aero-respiratory tract, gastrointestinal tract, liver and kidney, with limited presence within macrophages (CD68^+^ cells) and endothelial cells (CD105^+^ cells) of lung tissue (Fig. 1d-h). The S protein was only rarely detected in some of the SARS-CoV-2 PCR negative tissues tested, and not in post-mortem tissues from patients who did not have SARS-CoV-2 infection (data not shown). While SARS-CoV-2 S protein expression within lung alveolar epithelial cells was patchy in nature, consistent with possible aspiration of virus from the upper respiratory tract^13^, expression at non-pulmonary sites frequently revealed several well demarcated areas of confluent SARS-CoV-2 S protein expression within adjacent cells, surrounded by cells with no detectable protein (Fig. 1f-h). These ‘foci of infection’, with numerous affected cells adjacent to unaffected cells, are suggestive of cell-to-cell spread as reported in other coronavirus and respiratory viruses.^14,15^

Overall, we observed minimal evidence of acute inflammation in other organs (Fig. 1b-c). Background changes of chronic disease were common, reflecting pre-existing co-morbidities. Expected organ injury commensurate with severity of systemic illness was also present (e.g. renal acute tubular necrosis in mechanically ventilated patients). Detectable viral RNA in the kidney (n=4 detectable), liver (n=4) and gastrointestinal tract (n=7) was not associated with inflammation scores or with biochemical evidence of acute kidney injury, peak ALT measurement or enteric symptoms, respectively. No acute histological abnormalities were identified in the gastrointestinal tract or endocrine organs and no cases of myocarditis were identified despite frequent detection of viral RNA within these tissues. Importantly, the presence of viral protein within the kidney (n=4 assessed), intestine (n=3) and liver (n=2) were not associated with a localised inflammatory response adjacent to the infected cells (Fig. 1f-h).

### Pulmonary inflammation and relationship to SARS-CoV-2

The discrepancy between viral presence and inflammation at extra-pulmonary sites was also evident within the lung (Fig. 2a). The extent of viral RNA presence within the lung was not clearly associated with pulmonary inflammatory changes within our cohort. Diffuse alveolar damage (DAD) and bronchopneumonia were both observed in sections of lung with and without detectable virus. Similarly, virus could be detected in the absence of inflammation. For example, Patient I had viral RNA in all five lung lobes but histological examination demonstrated only mild, focal interstitial inflammation with no features of DAD; patient G had features of widespread bronchopneumonia in the context of widely detectable viral nucleic acid but, away from areas of bronchopneumonia, had no evidence of alveolar damage (Fig. 2a).

**Figure 2.**
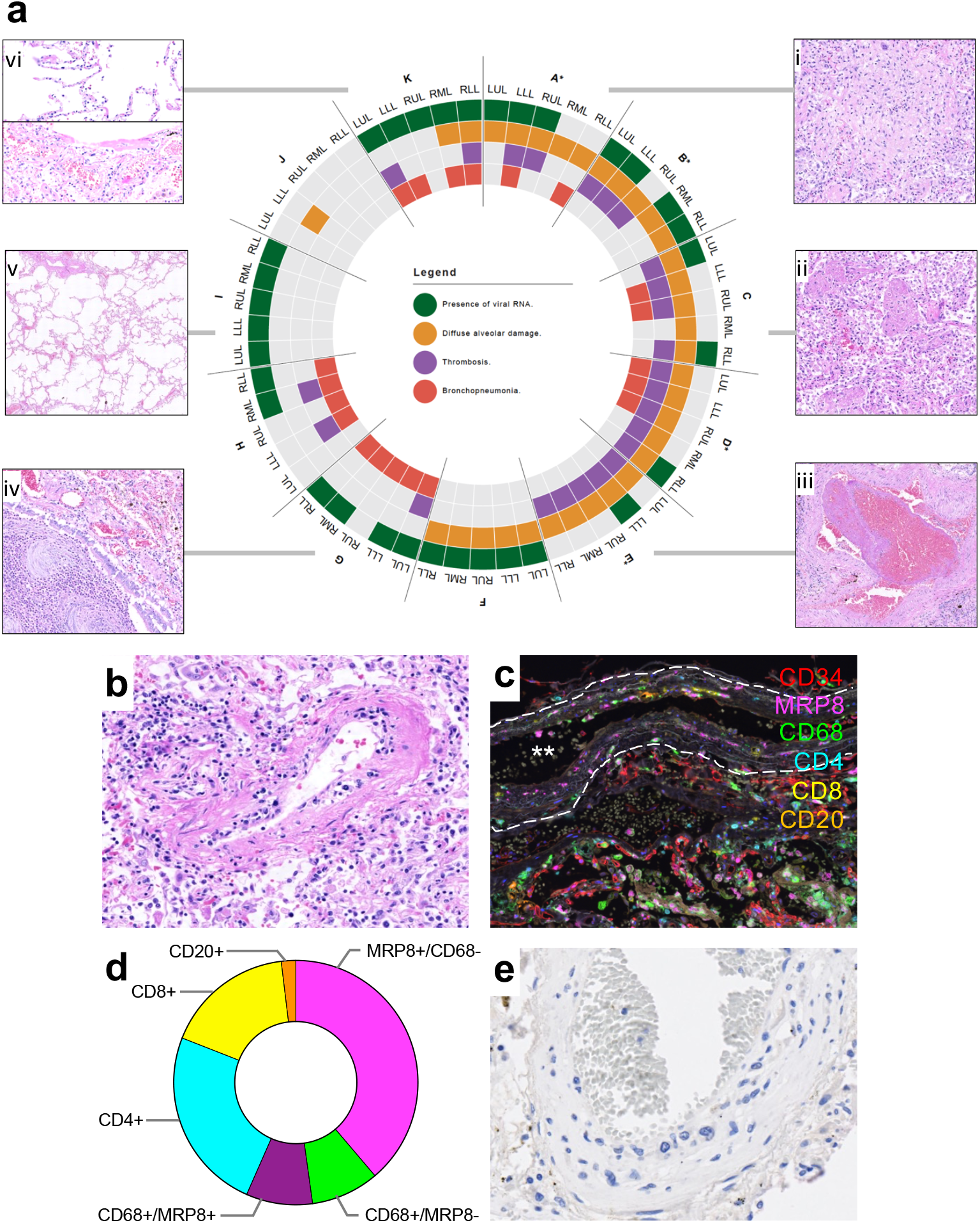
Delineating pulmonary injury and vascular involvement in fatal Covid-19. Detailed spatial evaluation of lung injury and key pathological abnormalities were determined within each lobe of lung for each patient and compared with the presence or absence of SARS-CoV-2 viral RNA by multiplex PCR (B; *denotes mechanical ventilation, n=11). Representative images of organising (i) and exudative (ii) diffuse alveolar damage, pulmonary thrombus (iii), bronchopneumonia (iv), uninflamed lung (v) and variable inflammation within the lung (vi). In four individuals frequent mononuclear cell vasculitis was seen within the pulmonary vasculature (C) with multiplex immunofluorescence defining vascular endothelium (CD34) and immune cell populations (CD4, CD8 (T cells); CD20 (B cells); CD68 (macrophages); MRP8 (neutrophils and myeloid lineage cells)) demonstrating MRP8 immunopositive mononuclear cells to be the predominant cell type (D: representative image, white stars denotes vessel lumen and white dashed line denotes elastic lamina; E: analysis of 50 arteries/arterioles from two selected patients quantifying cell types). SARS-CoV-2 S protein was not identified by immunohistochemical staining within endothelial cells of 40 inflamed vessels (F: representative image).

Consistent with recent reports, pulmonary thrombi were present in multiple patients (8/11; small vessel only n=1, large vessel only n=2, large and small vessel n=5) (Fig. 2a). In addition, a patchy but striking mononuclear cell vasculitis predominantly affecting intima of small/medium sized pulmonary arteries was observed in 4/11 cases (Fig. 2b). This inflammatory infiltrate in the affected pulmonary arteries was characterised in two patients (A&C) by H&E and multiplex immunofluorescence. MRP8^+^ mononuclear cells were demonstrated to be the predominant infiltrating population accompanied by a mixed population of CD4^+^ and CD8^+^ T cells and macrophages (Fig. 2c-d). Inspection of 40 inflamed vessels from the same patients did not identify SARS-CoV-2 S protein within the surface endothelium (Fig. 2e). No vasculitis was evident in any of the other organs studied.

Increased CD8^+^ T cells and reduced resident lung macrophages have recently been reported using single cell transcriptomics on bronchoalveolar lavage fluid (BALF) cells^16^. However, this approach risks underestimating pathophysiological and immune changes within the non-luminal pulmonary compartment. To complement these existing data, multiplex immunophenotyping was undertaken on whole lung tissue (Fig. 3a-f). Our analysis revealed that the greatest increase in immune cells were predominantly within parenchymal regions rather than vascular/perivascular areas (Fig. 3g). This showed that the largest relative increases were within the mononuclear phagocyte compartment (CD68^+^/MRP8^−^ macrophages, then CD68^+^/MRP8^+^ monocytic cells) followed by CD8^+^ then CD4^+^ T cells. Smaller increases in CD20^+^ cells and MRP8^+^/CD68^−^ neutrophils were also observed.

**Figure 3.**
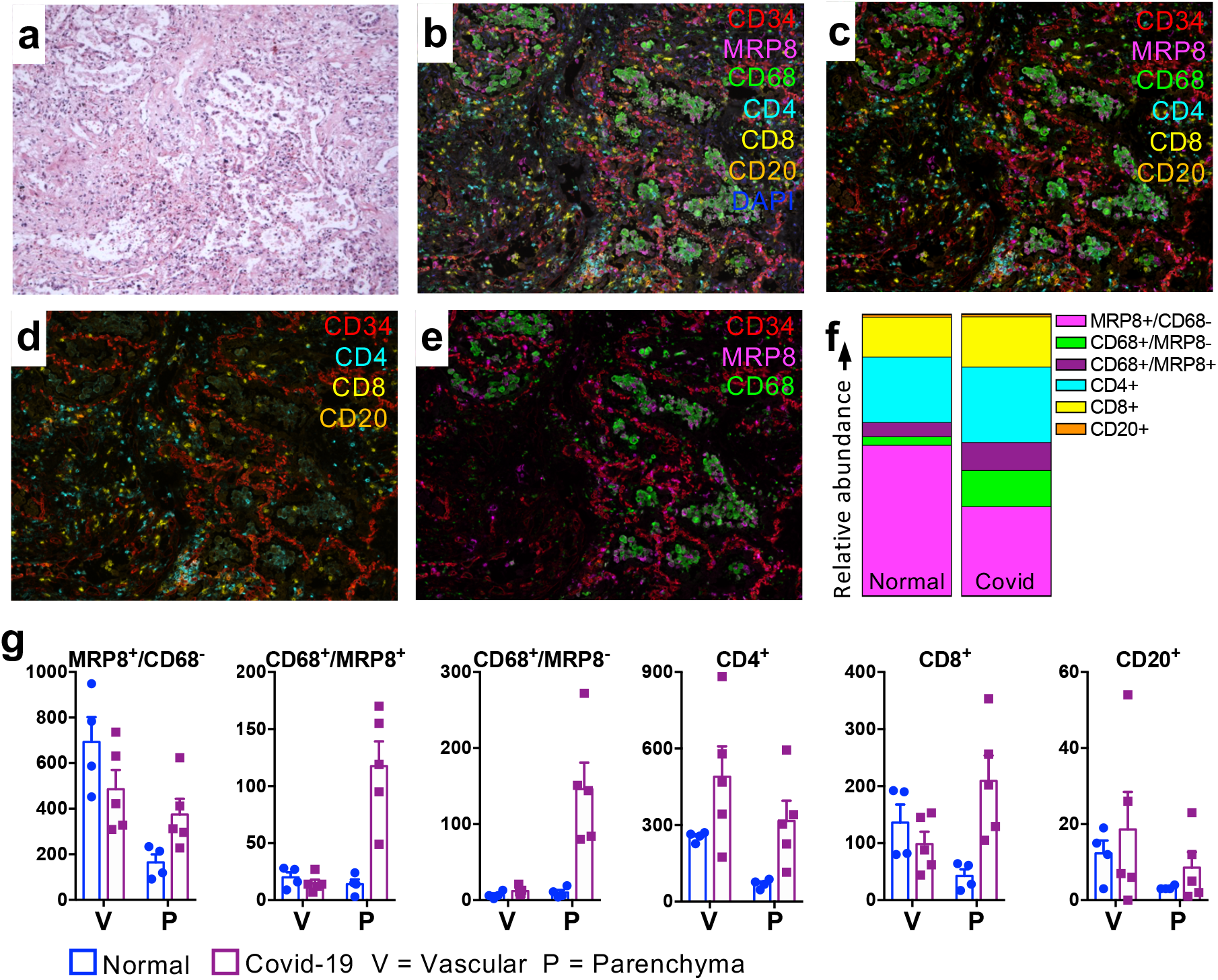
Pulmonary tissue immune response to fatal Covid-19. Regions of interest were defined by histological examination of lung tissue to identify areas of diffuse alveolar damage (DAD) in tissue from five patients (representative image, A). Corresponding multiplex immunofluorescence defined vascular endothelium (CD34) relative to immune cell populations: CD4, CD8 (T cells); CD20 (B cells); CD68 (macrophages); MRP8 (neutrophils and myeloid lineage cells) with (B) and without (C) autofluorescence. Separate cell populations are highlighted in panels D-F. Immune cell populations were quantified and spatially stratified (G) into vascular/perivascular and parenchymal regions, with relative abundance of cell types compared between Covid-19 (n=5) and normal, uninflamed lung from patients undergoing lung cancer resection (n=4) (H).

### Reticulo-endothelial system responses in fatal Covid-19

All cases showed a severe stereotyped pattern of pathological change regardless of viral RNA presence within the lymph node or spleen (Fig. 4a). Within the bone marrow, erythroid dysplasia, plasma cell excess with morphological atypia and iron storage abnormalities were identified (Fig. 4b-c). A marked increase in the number of plasma cells (10% or more) was seen in 7/8 bone marrow trephines but these plasma cells had a normal phenotype, being negative for CD56 and cyclin D1, and were polytypic with light chain immunohistochemistry. Iron laden macrophages were seen in 7/8 samples and associated with abundant iron storage on Perl’s stain. Although infrequent (1-2/1,000 cells), haemophagocytosis of erythroid and/or myeloid precursors was present in bone marrow in three cases. In mediastinal lymph nodes, marked reactive plasmacytosis of CD38^+^/MUM1^+^ and weakly CD138+ cells were seen in the paracortex and medulla, again exhibiting a degree of nuclear polymorphism. In the spleen, white pulp atrophy was common (4/7) similar to post-mortem observations in fatal SARS^17,18^. Splenic red pulp was congested and, in all cases, contained an increased number of plasma cells with similar features to those observed in mediastinal nodes.

**Figure 4.**
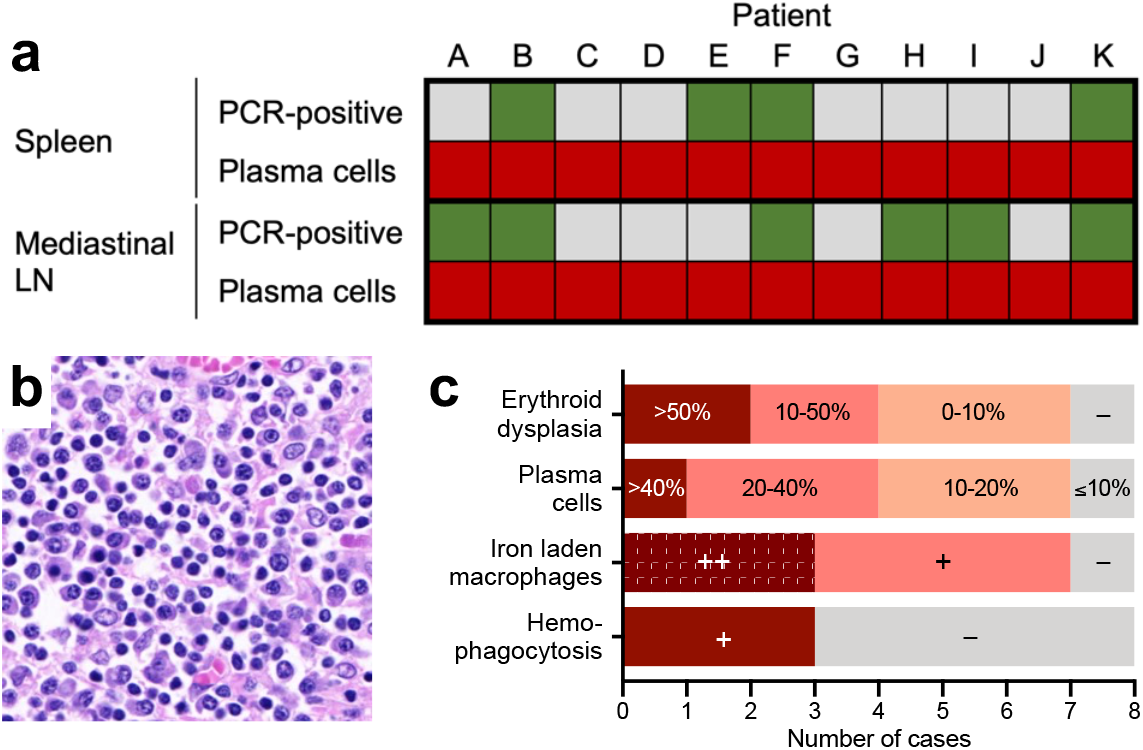
Reticulo-endothelial response in fatal Covid-19. Mismatch between consistent plasma cell abnormalities in spleen and mediastinal lymph node (LN) (red) and detection of SARS-CoV-2 by PCR (A: green: PCR positive; grey: PCR negative) Numerous plasma cells were present throughout the lung and reticuloendothelial system (B: representative image from mediastinal lymph node). Key pathological abnormalities within bone marrow aspirates included erythroid dysplasia, iron-laden macrophages and haemophagocytosis; plasma cells were confirmed by immunohistochemical staining and quantified in bone marrow trephines (C).

## DISCUSSION

The present study shows that fatal Covid-19 is associated with variable but widespread distribution of viral RNA and protein but with a discordant inflammatory response to local viral presence, both between and within tissues, demonstrating tissue-specific tolerance of SARS-CoV-2. The overall picture suggests an aberrant immune response, principally involving the lung and reticuloendothelial system, that is not clearly topologically associated with the virus.

If organ injury is primarily collateral damage to an appropriate local inflammatory response against SARS-CoV-2, it would be expected to have a temporal and spatial association with the presence of the virus. We have observed the opposite and find that viral products (both RNA and protein) are present in numerous tissues at time of death in fatal Covid-19, persisting up to 42 days after illness onset. The presence of viral RNA within the kidney, intestine and liver was not associated with histological evidence of organ injury or inflammation. By spatially resolving viral presence we confirmed that in extra-pulmonary tissues, cells containing the S protein did not have an adjacent localised inflammatory response. This is consistent with strains of avian coronavirus which can replicate in the gut without causing macroscopic or histological changes.^19^

Consistent with these extra-pulmonary observations, there is not a consistent association between viral presence and either the presence or nature of the inflammatory response within the lung in fatal Covid-19. Within our cohort, we report (i) patients with detectable pulmonary virus and DAD, (ii) virus and bronchopneumonia without DAD, and (iii) widespread virus without inflammation. Consistent with existing literature, small and large pulmonary vessel thrombi were common in our series.^20,21^ Thrombi in pulmonary vessels have also been reported in fatal cases of SARS^17,22^, H1N1pdm09 influenza A virus infection,^23,24^ and ARDS more generally, but the frequency in Covid-19 appears nearly a log order higher compared to influenza and may be due to distinct endothelial injury pathways^20^.

Here we report a high frequency of pulmonary vasculitis characterised by a predominantly MRP8^+^ mononuclear cell (monocyte/myeloid) infiltration into the vessel wall. The vasculitis was not overtly associated with local endothelial viral S protein expression although its presence was identified within a small number CD105^+^ endothelial cells in other vessels within the lung. This observation validates the drive to understand the immune microenvironment at a whole lung level. Building on the observation of increased CD8^+^ T cells and reduced resident lung macrophages in BALF^16^, we describe a relative increase in mononuclear phagocytes and to a lesser extent CD4^+^ and CD8^+^ T cells, within the non-luminal pulmonary compartment. Furthermore, iron laden macrophages were observed within bone marrow in all but one patient, despite the absence of typical causes of secondary iron overload (transfusion, haemolysis), consistent with the observation that circulating ferritin correlates with adverse outcomes^25^. In human immunodeficiency virus and hepatitis C virus infection, iron overload is associated with poor prognosis, with evidence that viral infection itself may enhance macrophage iron loading, further suggesting that iron overload is an aberrant response deleterious to the host in Covid-19^26^. It will be important to clarify whether these macrophages, within inflamed pulmonary vessels, lung parenchyma and reticulo endothelial tissues, have an anti-viral or tissue repair role, or whether being activated as part of the wider immune response to virus they are themselves promoting vascular and tissue injury. The implications for opposing strategies to either boost or inhibit macrophage function are obvious, and necessitate urgent further investigation.

Plasma cell abnormalities in the reticuloendothelial system and lung provided further evidence of an aberrant host response in fatal Covid-19. While plasma cell expansion is expected to ensure production of antibody in the context of acute infections, the levels seen in our study were extremely marked. Plasma cells exhibited morphologic atypia but displayed a reactive, polytypic phenotype. To some extent this correlates with peripheral blood findings in patients with Covid-19 where CD4^+^ and CD8^+^ T-cell depletion is characteristic but B-cell numbers are maintained, with higher B-cell numbers reported in severe cases^27,28^. The plasma cells in our study were generally MUM1^+^ and CD38^+^ but CD138 (syndecan) low/negative raising the possibility that these are short-lived plasma cells or are at a transitional or arrested stage of development^29^. In addition to macrophage behaviour and iron accumulation, this identifies plasma cells as a priority for future investigation of therapeutic targets.

Taken together, the present data provide comprehensive clinical, viral and inflammatory profiling of fatal Covid-19. These highlight, for the first time, the discrepancy between the presence of SARS-CoV-2 and tissue inflammation. Given the recent discovery that immunosuppression with dexamethasone prevents death in severe Covid-19, our observations support virus-independent immunopathology being one of the primary mechanisms underlying fatal Covid-19. This suggests that better understanding of non-injurious, organ specific viral tolerance mechanisms and targeting of the dysregulated immune response merits further investigation within the broader context of management of Covid-19.

## Data Availability

All data are available on request from the corresponding authors

## METHODS

### Post-mortem examinations

Post-mortem examinations were conducted in a biosafety level three (BSL3) post-mortem facility on patients with pre-mortem PCR-confirmed SARS-CoV-2 infection and evidence of lower respiratory tract disease at a median of 19·3 hours after death (interquartile range 4·6– 20·2). 37 tissue sites were sampled for histology and RNA analyses including 23 from the respiratory tract. Samples were fixed in formalin or treated with TRIzol, snap frozen and stored at −80°C. Ethical approval was granted by the East of Scotland Research Ethics Service (16/ES/0084). The patients had a mean age of 77 years (range 64–97), 10/11 were male, and four received invasive mechanical ventilation during their hospital admission.

### Tissue histology and immunofluorescence

Formalin-fixed paraffin-embedded (FFPE) tissue blocks were processed, haematoxylin and eosin stained and reviewed by specialist histopathologists. For immunophenotyping, multiplexed immunofluorescence on de-paraffinised rehydrated FFPE slides was performed using combinations of primary antibodies against CD34, CD68, MRP8, CD4, CD8 and CD20, labelled with TSA-conjugated fluorophores, with antibody removal between steps. Images were captured using a Vectra Polaris slide scanner (Akoya Biociences).

### Viral RNA and protein detection

Total RNA was extracted at BSL3 from homogenised TRIzol treated tissue. Samples were DNAse treated and cDNA synthesised before amplification of SARS-CoV-2 by the ARTIC Network protocol using the multiplexed primer scheme version three. Purified PCR products were processed, sequenced and analysed as per the appendix. Post-mortem interval was not associated with the number of tissue samples that were SARS-CoV-2 PCR positive post-mortem. De-paraffinised, rehydrated FFPE slides were examined for presence of SARS-CoV-2 using immunohistochemistry to detect SARS-CoV-2 spike (S) protein and cell specific markers (CD68, AE1/3 and CD105), to detect viral presence.

## ACKNOWLEDGEMENTS

ICECAP research depends on the generosity of donors and their families who provide the valuable gift of tissue after death.

ICECAP receives funding and support from The Chief Scientist Office (RARC-19 Funding Call, ‘Inflammation in Covid-19: Exploration of Critical Aspects of Pathogenesis; COV/EDI/20/10’ to D.A.D, C.D.L, C.D.R, J.K.B and D.J.H), LifeArc (through the University of Edinburgh STOPCOVID funding award, to K.D, D.A.D, C.D.L) and Medical Research Scotland (CVG-1722-2020 to D.A.D, C.D.L, C.D.R, J.K.B and D.J.H). C.D.L is a Wellcome Trust Clinical Career Development Fellow (206566/Z/17/Z). J.K.B. and C.D.R. are supported by the Medical Research Council (grant MC_PC_19059) as part of the ISARIC Coronavirus Clinical Characterisation Consortium (ISARIC-4C). D.J.H, I.H.U and M.E are supported by iCAIRD (Industrial Centre for Artificial Intelligence Research in Digital Diagnostics). S.P. is supported by Kidney Research UK and Giulia by The Melville Trust for the Cure & Care of Cancer. Identification of SARS-CoV-2 and sequencing work was supported by the United States Food and Drug Administration grant number HHSF223201510104C ‘Ebola Virus Disease: correlates of protection, determinants of outcome and clinical management’ amended to incorporate urgent COVID-19 studies awarded to J.A.H. R.P.-R. is directly supported by the Medical Research Council Discovery Medicine North Doctoral Training Partnership. The group of J.A.H. is supported by the National Institute for Health Research Health Protection Research Unit (NIHR HPRU) in Emerging and Zoonotic Infections at University of Liverpool in partnership with Public Health England (PHE), in collaboration with Liverpool School of Tropical Medicine and the University of Oxford. J.A.H. is also funded by the Centre of Excellence in Infectious Diseases Research (CEIDR) and the Alder Hey Charity.

## AUTHOR CONTRIBUTIONS

DAD, CDL, CDR, JKB, DJH, JAH, CS: Conceptualization, Methodology, Validation, Formal analysis, Investigation, Data Curation, Writing – Original Draft, Writing – Review & Editing, Visualization, Supervision, Project administration, Funding acquisition. IHU, ME, SDA, RP-R, TM, CEBL, GT, CSH, NPR, VLC, AJD, WAQ, RB, COCB, SP, WAW: Investigation, Formal analysis, Writing – Review & Editing. KD, AMA, NNG: Resources. PMDP: Visualization.

